# Household Secondary Attack Rate of COVID-19 and Associated Determinants

**DOI:** 10.1101/2020.04.11.20056010

**Authors:** Qin-Long Jing, Ming-Jin Liu, Jun Yuan, Zhou-Bin Zhang, An-Ran Zhang, Natalie E. Dean, Lei Luo, Mengmeng Ma, Ira Longini, Eben Kenah, Ying Lu, Yu Ma, Neda Jalali, Li-Qun Fang, Zhi-Cong Yang, Yang Yang

## Abstract

**Background:** As of April 2, 2020, the global reported number of COVID-19 cases has crossed over 1 million with more than 55,000 deaths. The household transmissibility of SARS-CoV-2, the causative pathogen, remains elusive.

**Methods:** Based on a comprehensive contact-tracing dataset from Guangzhou, we estimated both the population-level effective reproductive number and individual-level secondary attack rate (SAR) in the household setting. We assessed age effects on transmissibility and the infectivity of COVID-19 cases during their incubation period.

**Results:** A total of 195 unrelated clusters with 212 primary cases, 137 nonprimary (secondary or tertiary) cases and 1938 uninfected close contacts were traced. We estimated the household SAR to be 13.8% (95% CI: 11.1-17.0%) if household contacts are defined as all close relatives and 19.3% (95% CI: 15.5-23.9%) if household contacts only include those at the same residential address as the cases, assuming a mean incubation period of 4 days and a maximum infectious period of 13 days. The odds of infection among children (<20 years old) was only 0.26 (95% CI: 0.13-0.54) times of that among the elderly (≥60 years old). There was no gender difference in the risk of infection. COVID-19 cases were at least as infectious during their incubation period as during their illness. On average, a COVID-19 case infected 0.48 (95% CI: 0.39-0.58) close contacts. Had isolation not been implemented, this number increases to 0.62 (95% CI: 0.51-0.75). The effective reproductive number in Guangzhou dropped from above 1 to below 0.5 in about 1 week.

**Conclusion:** SARS-CoV-2 is more transmissible in households than SARS-CoV and MERS-CoV, and the elderly ≥60 years old are the most vulnerable to household transmission. Case finding and isolation alone may be inadequate to contain the pandemic and need to be used in conjunction with heightened restriction of human movement as implemented in Guangzhou.

## Introduction

The ongoing pandemic of the coronavirus disease 2019 (COVID-19), caused by the novel coronavirus SARS-CoV-2, has now affected 181 countries worldwide. As of April 2, 2020, there were more than 1 million reported cases and over 55,000 deaths.^1^ The elderly and individuals with chronic conditions such as diabetes and cardiopulmonary disease are most vulnerable to severe disease and death. Efficient transmission via droplets and fomites is potentially supplemented by other transmission routes such as aerosol and fecal contamination.^2,3^ Evidence is emerging that pre-symptomatic or asymptomatic carriers can transmit the virus.^5,6^ It is suspected that within-household transmission might have contributed substantially to the continued rise in cases in China even after the introduction of nationally enforced restrictions on human movement.^7,8^ Home isolation/quarantine of people with an exposure history or mild symptoms is frequently recommended as a disease control measure in countries with COVID-19 outbreaks, but such restrictions likely have limited or no effect on family transmission.

Thus far, transmissibility of the disease has primarily been assessed either at the population level using mathematical models or at the individual level in synthetic populations using agent-based models.^9-11^ Transmissibility within households or through other types of close contact remains under-investigated, despite the importance of these social interactions in shaping the overall dynamics of disease spread and in determining the effectiveness of mitigation strategies.^12^ Obtained via tracing, contact data provide the most accurate information about human-to-human transmissibility of any infectious pathogen, because transmissibility can be assessed conditioning on exposure. Currently available estimates for the secondary attack rate (SAR) of SARS-CoV-2 were based on contact-tracing data of hundreds of cases in Shenzhen and Guangzhou, Guangdong Province in southern China as well as 10 cases in the United States.^13-15^ These estimate are proportions of confirmed infections among all traced contacts, not accounting for heterogeneity in exposure level, possibility of transmission among contacts themselves and infection risks from untraced contact or fomites. We refer to these proportions as the non-primary attack rate rather than SAR, as the infections among contacts are not necessarily secondary.

Applying a statistical transmission model to the contact-tracing data from Guangzhou, we estimated the SAR of SARS-CoV-2 among household and non-household close contacts. We assessed the effects of age and gender on the infectiousness of COVID-19 cases and susceptibility of their close contacts. In addition, we evaluated the relative infectiousness of COVID-19 cases during the incubation period compared to the period of illness.

## Methods

### Case definition

Case definitions (Appendix Sec. 1.1) are in accordance with the Control and Prevention Plan for the Novel Coronavirus Pneumonia Disease (v5) issued by the Chinese Center for Disease Control and Prevention (China CDC). Briefly, a suspected COVID-19 case is defined as a patient meeting ≥1 epidemiological criteria and ≥2 clinical criteria as outlined in the appendix. A confirmed case is defined as a suspected case with positive detection of SARS-CoV-2 nucleic acid by real-time RT-PCR or viral genes that are highly homologous to SARS-CoV-2 by sequencing using respiratory or blood specimens. An asymptomatic infection is an individual with laboratory confirmation but without clinical signs, mainly found by outbreak investigation and contact tracing.

### Epidemiological investigation and contact-tracing

Epidemiological investigations were conducted by county-level CDC offices within 24 hours after a suspected or confirmed case or an asymptomatic infection was reported. For each suspected or confirmed case, demographic, clinical, diagnostic and occupational data, baseline health conditions, clinical samples and laboratory test results, and exposure history in the 14 days prior to symptom onset were recorded using a standardized investigation form. Typically, a suspected case would be changed to a confirmed case or removed from the surveillance system when laboratory test results became available. A close contact is defined as an individual who had unprotected close contact (within 1 meter) with a confirmed case or an asymptomatic infection within 2 days before their symptom onset or sample collection, including but not limited to household members, care givers and individuals in the same workplace, classroom, ward or transportation vehicle. Close contacts were quarantined at an observatory place or at home and followed for 14 days, according to the Temporary Guidelines for Investigation and Management of Close Contacts of Novel Coronavirus Pneumonia Patients issued by China CDC.

In our analyses, individuals who are linked by contact tracing are considered as a cluster. Given a cluster, we define cases with symptom onset on either the earliest onset day or the following day as primary cases. If a cluster had only asymptomatic infections, primary cases are determined by specimen collection dates. Individuals who are either family members, or close relatives, such as parents and parents-in-law, or live in the same residential address are considered as household contacts of each other. In sensitivity analyses, we also restrict household contacts to individuals living at the same address.

### Statistical Analysis

Standard nonparametric tests such as Fisher’s exact test were used to compare characteristics between demographic groups in R.^16^ The spatial distribution of clusters was mapped at the community level using ArcGIS (Environmental Systems Research Institute, Redlands, California), with a directed graph indicating potential transmission chains. We estimated effective reproductive numbers (*R*_*t*_) based on the contact tracing data (Appendix Sec. 1.2). Due to the uncertainty in the transmission relationship, we explore three scenarios defined by the following assumptions: (1) all imported cases (with travel or living history in Wuhan or Hubei Province 14 days before symptom onset) were primary cases, and all nonprimary cases were infected by primary cases in the same cluster; (2) in addition to (1), local primary cases might have been infected by earlier primary cases in other clusters; and (3) in addition to (2), imported non-primary cases were not considered as primary cases.

A chain-binomial statistical model was used to estimate SAR and local reproductive number (Appendix Sec. 1.3).^17^ Only confirmed infections (symptomatic or asymptomatic) and their contacts were included in the analysis. the Expectation and maximization (EM) approach was used to account for uncertainty in the infection time of asymptomatic infections (Appendix Sec. 1.3). Possible distributions of the incubation period and the infectious period were derived from the literature and our previous work on a separate contact-tracing database (Appendix Sec. 1.4).^18-20^

Briefly, this model estimates the probabilities, denoted by *p*_1_ and *p*_2_, of viral transmission from an infectious household contact and from an infectious non-household contact (e.g., friends, co-workers, passengers, etc.), respectively, to a susceptible person per daily contact. In addition, each susceptible person is subject to a constant daily probability, *b*, of being infected by an unspecified external source, which accounts for untraced contacts and fomites. Suppose that the infectivity of a COVID-19 case differs between the incubation period and the post-illness-onset period (referred to as the illness period hereinafter). We use (*D*_*min*_, *D*_*max*_) to represent the whole infectious period with the symptom onset day set as day 0. We then model the effective daily transmission probability as 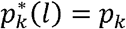 on days *D*_*min*_ : ≤ *l* < 0 (incubation period) and 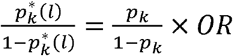 on days 0 ≤ *l* < D_max_ (illness period), where *OR* is the odds ratio measuring the relative infectivity of the illness period vs. the incubation period. The SAR is defined as

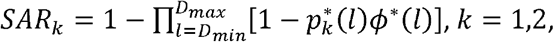

where *ϕ*^*^(*l*) specifies the relative infectivity level on the *l*_*th*_ day of the infectious period based on prior studies,^20^ peaking near the time of symptom onset. We define the local reproductive number as the average number of infections a symptomatic case can generate via household and non-household contacts,

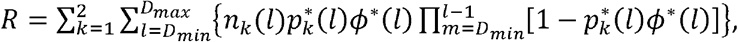

where *n*_1_(*l*) and *n*_2_(*l*) are the average numbers of household and non-household contacts per primary case on day *l* (Appendix Sec. 1.5).

We assessed the effect of age and gender on susceptibility or infectivity by regressing transmission probabilities on these demographic characteristics. Finally, we evaluated the goodness-of-fit of the model under different settings of the natural history of disease (Appendix Sec. 1.6).

### Ethics statement

Data were collected as part of an continuing public health response required by the national Health Commission of China, and hence informed consent was waived. Analysis of deidentified data was considered not to be human subjects research after consultation with the University of Florida IRB. All analyses of personally identifiable data were conducted at the Guangzhou CDC.

## Results

By February 17, 2020, a total of 349 lab-confirmed cases were reported to Guangzhou Municipal CDC, forming 195 unrelated clusters with 212 primary cases and 137 nonprimary (secondary or tertiary) cases. The size of clusters ranges from 1 to 274, with a median size of 6 and an inter-quartile range (IQR) of (4, 10). The majority (65%) of clusters had no secondary cases.

The majority of cases were adults aged 20-59 years (Table 1). The majority of primary cases (171, 81%) and more than half of non-primary cases (77, 56%) had recent travel or living history outside Guangzhou (referred to as imported cases hereinafter), and 89% of these imported cases were related to Hubei Province. Among the 155 imported primary cases with known arrival dates, 123 (79%) arrived at Guangzhou on or before the complete lockdown of Wuhan (Jan. 24). The overall non-primary attack rates are 12.6% and 3.06% among household and non-household contacts, respectively. Within households, the non-primary attack rate was much lower in contacts <20 years group, 5.26% (95% CI 2.43, 9.76%), as compared to 13.72% (95% CI 10.68, 17.24%) and 17.69% (95% CI: 11.89, 24.83%) in 20-59 year olds and ≥60 year olds, respectively (p-values <0.005). A similar age trend was observed among non-household contacts, but the differences were not of statistical significance. There was no gender difference in the non-primary attack rates, for both inside and outside the household.

**Table 1.** Demographic compositions of the study population stratified by case type (primary, nonprimary and non-case) and contact type (household and non-household). Percentages are presented in parentheses. Non-primary incidence is calculated as the number of non-primary cases divided by the sum of non-primary cases and non-cases.

| Demographic feature | Category | Primary cases | Nonprimary cases |  | Non-cases |  | Overall | Non-primary Attack Rate (%) |  |
| --- | --- | --- | --- | --- | --- | --- | --- | --- | --- |
|  |  |  | Household | Non-household | Household | Non-household |  | Household | Non-household |
| Age group | <20 | 10 ( 5) | 9 ( 9) | 1 (3) | 162 (24) | 72 ( 6) | 254 (11) | 5.26 (2.43, 9.76) | 1.37 (0.035, 7.4) |
|  | 20-59 | 142 (67) | 62 (64) | 30 (75) | 390 (58) | 950 (75) | 1574 (69) | 13.72 (10.68, 17.24) | 3.06 (2.07, 4.34) |
|  | ≥60 | 60 (28) | 26 (27) | 9 (22) | 121 (18) | 243 (19) | 459 (20) | 17.69 (11.89, 24.83) | 3.57 (1.65, 6.67) |
| Gender | Female | 105 (50) | 56 (58) | 20 (50) | 340 (51) | 617 (49) | 1138 (50) | 14.14 (10.86, 17.97) | 3.14 (1.93, 4.81) |
|  | Male | 107 (50) | 41 (42) | 20 (50) | 333 (49) | 648 (51) | 1149 (50) | 10.96 (7.98, 14.58) | 2.99 (1.84, 4.59) |
| Origin | Imported | 171 (81) | 50 (52) | 27 (67) |  |  |  |  |  |
|  | Local | 41 (19) | 47 (48) | 13 (33) |  |  |  |  |  |
| Total |  | 212 (100) | 97 (100) | 40 (100) | 673 (100) | 1265 (100) | 2287 (100) | 12.60 (10.34, 15.15) | 3.06 (2.2, 4.15) |

Most cases were found in the densely populated districts (56% of the total population in Guangzhou) including Yuexiu, Liwan, Haizhu, Tianhe and Baiyun (Figure 1). Three clusters with 5 or more secondary cases (not counting tertiary or further generations) were observed, one in each of Yuexiu, Haizhu and Baiyun districts, and all primary cases in these clusters were imported (Figure 1A-B). The longest transmission chain had three subsequent generations from the primary case, which occurred in Panyu district (Figure 1C). Five other clusters had two subsequent generations (Figure 1A-B). The reported residential locations of primary and non-primary cases within the same clusters are mostly identical, but non-household non-primary cases might live far from the primary cases. Most transmissions occurred between household members (Appendix Figure S1).

**Figure 1.**
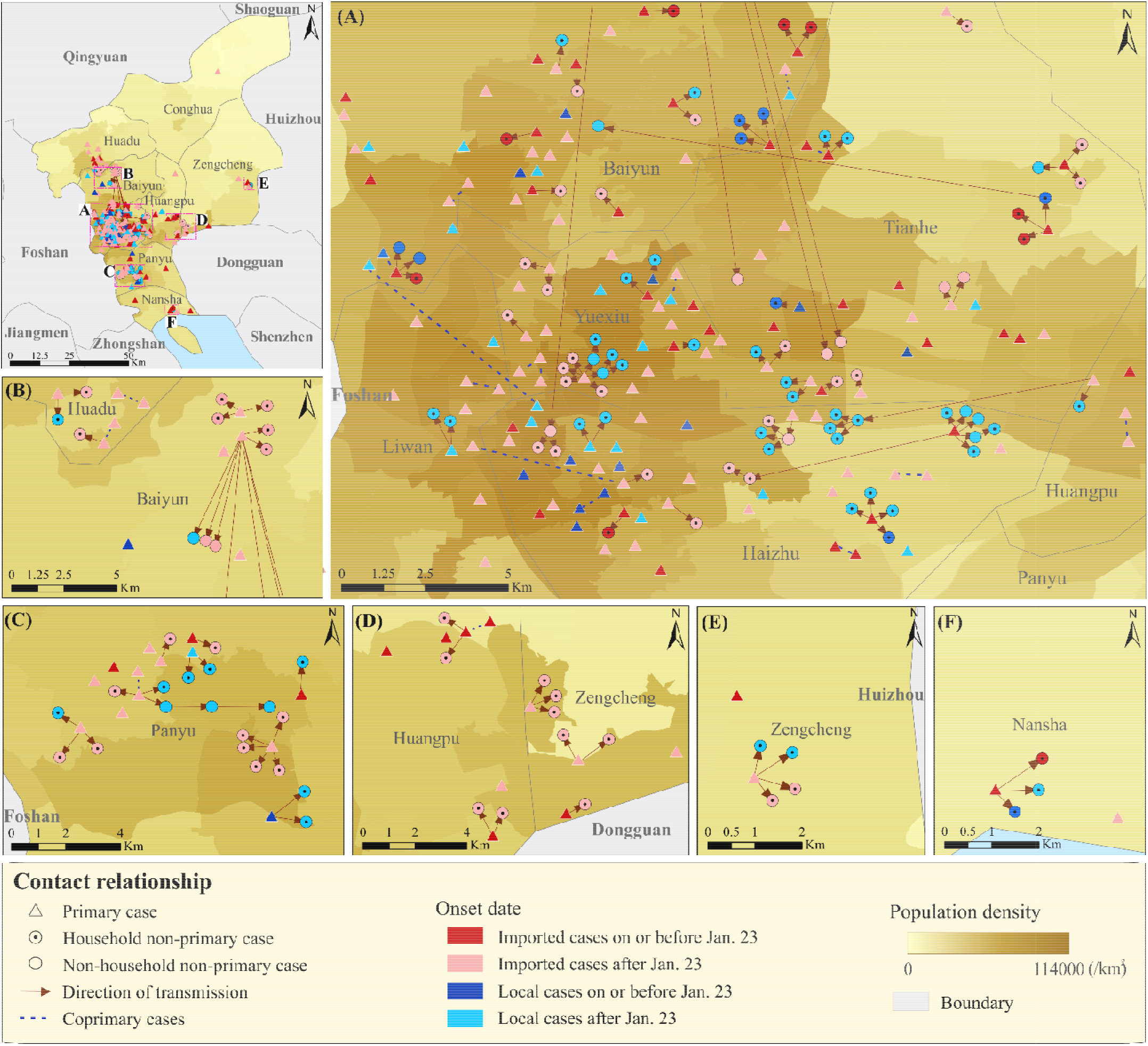
Spatial distribution of COVID-19 case clusters based on contact-tracing data from Guangzhou through February 17, 2020. The upper-left panel shows the overall distribution in Guangzhou, and panels A-F show the distribution in the sub-regions defined in the upper-left panel. Cases are considered as primary (triangles) if their symptom onset dates are the earliest or one day after the earliest in the cluster and as non-primary (circles) otherwise. Infected household contacts are represented by dotted circles. Non-infected contacts are not shown. Cases imported from outside Guangzhou are colored in red and local cases are colored in blue. Darker (lighter) color indicates symptom onset on or before (after) Jan. 23, 2002. Each arrow indicates potential transmission direction from a case with an earlier symptom onset to an infected contact with a later symptom onset. Co-primary cases are linked by unidirectional dashed lines. The displayed location of each case is jittered away from the actual residential address. Population densities at the township level are shown as the background.

Figure 2 shows the temporal trend of the COVID-19 epidemic in Guangzhou. The first imported primary case had symptom onset on Jan. 7, 2020, and this case arrived in Guangzhou on Jan. 13. The earliest local primary case had symptom onset on Jan. 16, by which time there were already at least 2 imported cases in Guangzhou. The number of imported cases peaked together with the epidemic in Guangzhou around Jan. 27, 4 days after the lockdown in Wuhan. After that, both case importation and the epidemic itself waned fast, and only sporadic cases were reported by the middle of February. The *R*_*t*_ started relatively high at the early phase, reaching 1.3 in scenario (3), barely 1.1 in scenarios (2) and only 0.7 in scenario (1). *R*_*t*_ quickly declined after the peak to below 0.5 by Jan. 27 for all scenarios, likely reflecting the tightening of control measures in Guangzhou.

**Figure 2.**
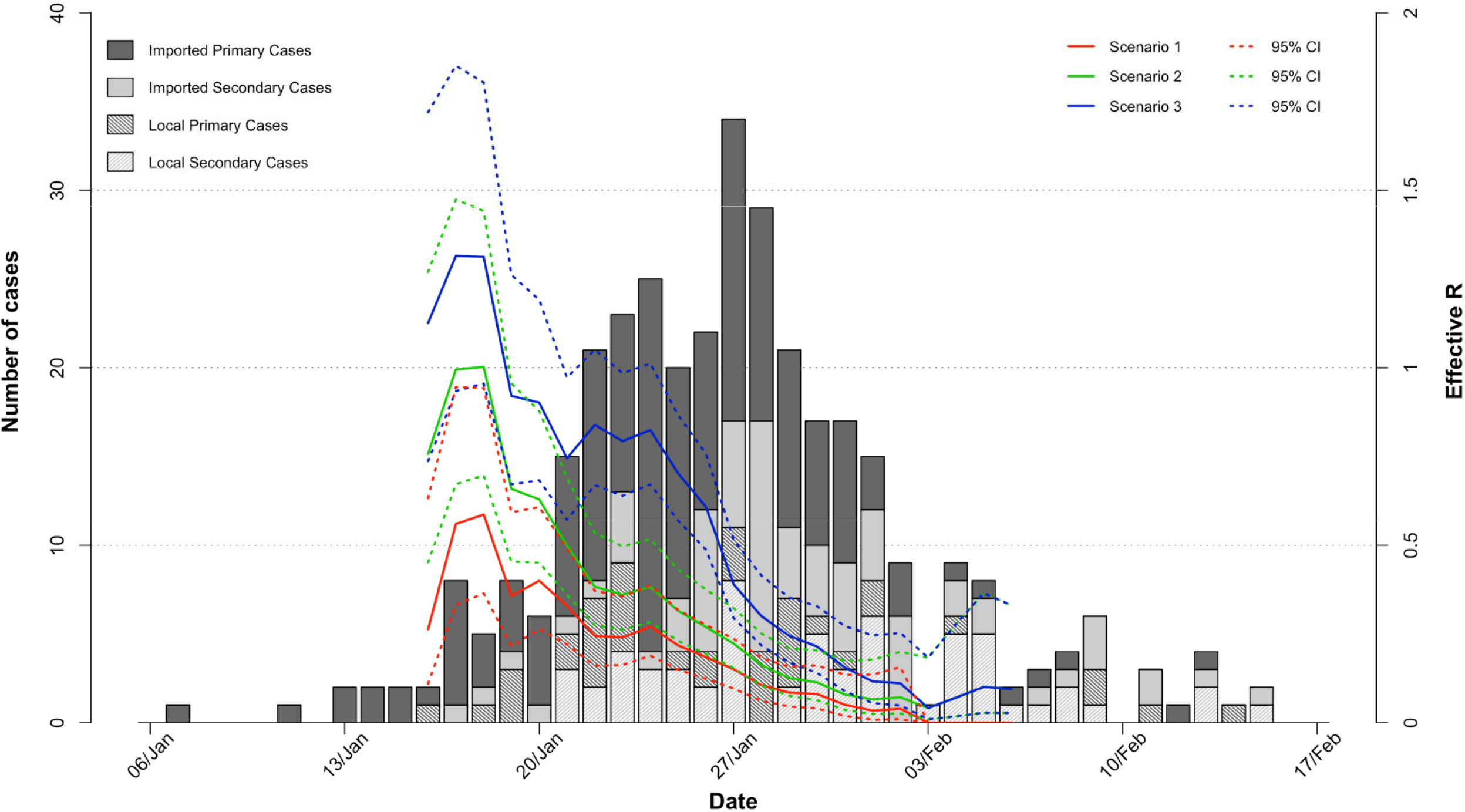
Epidemic curve based on symptom onset dates of COVID-19 cases in Guangzhou from Jan. 6 to Feb. 18, 2020. Cases are stratified by imported vs. local and primary vs. secondary and shaded correspondingly. Here all non-primary cases are considered as secondary cases. Estimated effective reproductive numbers *R*_*t*_ are shown for three scenarios corresponding to assumptions about the transmission relationship between different types of cases (see methods, statistical analysis).

After excluding 12 clusters with only primary cases but no contact, we included in the transmission analysis 183 clusters with a total of 335 cases (329 symptomatic and 6 asymptomatic) and 1938 noninfected contacts. We estimated the household and non-household SARs for the combinations of mean incubation period of 4-7 days and maximum infectious period of 13, 16 and 19 days. Assessment of the goodness-of-fit for these settings suggests that all models fits the data reasonably well, with model predicted infection numbers falling largely in line with the observed values. A mean incubation period of 4 days and a maximum infectious period of 13 days yielded the smallest difference between observed and model-fitted numbers (Appendix Figure S3). Consequently, we reported the estimates associate with this setting as the primary results.

When household contact is defined based on close relatives, we estimate the household SAR as 13.8% (95% CI: 11.1-17.0%) and the non-household SAR as 7.1% (95% CI: 4.7-10.6%), had there been no case isolation. A longer incubation periods is associated with a slightly lower, whereas a longer infectious period is associated with a slightly higher, SAR estimate (Table 2; Appendix Table S3). The household SAR varies from 12% to 17%, and the non-household SAR varied from 6% to 9%. The local R based on observed contact frequencies of primary cases was estimated to be 0.48 (0.39, 0.58), which is insensitive to the setting for the incubation and infectious periods. In other words, a typical case infected 0.48 individuals on average in Guangzhou, implying an inefficient transmission of the disease under the control measures. The projected local R, had there been no quarantine of cases, was estimated as 0.62 (0.51-0.75). High estimates of projected local R were associated with short incubation and longer infectious periods (Appendix Table S3). The daily transmission probability during the incubation period tends to be similar to that during the illness period (Table 3), with an estimated odds ratio (OR) of 1.13 (0.59-2.18); however, the difference is much wider when longer incubation periods were assumed (Table3; Appendix Table S4-S5).

**Table 2.** Model-based estimates (and 95% confidence intervals) of secondary attack rates (SAR) among household and non-household contacts, and model-based estimates of local reproductive number (local R) with and without quarantine. Estimates are reported using two different definitions of household contact (close relatives, or only individuals sharing the same residential address) and for selected settings of the natural history of disease. This model is not adjusted for age group.

| Definition of household contact | Parameter | Setting | Mean incubation period = 4 days |  | Mean incubation period = 7 days |  |
| --- | --- | --- | --- | --- | --- | --- |
|  |  |  | Max infectious period = 13 days | Max infectious period = 19 days | Max infectious period = 13 days | Max infectious period = 19 days |
| Close relatives | SAR (%) | Household | 13.8 (11.1, 17.0) | 16.8 (13.3, 21.1) | 11.7 (9.4, 14.5) | 12.8 (9.9, 16.3) |
|  |  | Non-household | 7.1 (4.7, 10.6) | 9.0 (5.8, 13.6) | 6.2 (4.1, 9.2) | 6.8 (4.4, 10.4) |
|  | Local R | With quarantine | 0.48 (0.39, 0.58) | 0.48 (0.39, 0.60) | 0.48 (0.39, 0.59) | 0.49 (0.38, 0.62) |
|  |  | No quarantine | 0.62 (0.51, 0.75) | 0.76 (0.61, 0.95) | 0.53 (0.43, 0.65) | 0.58 (0.45, 0.74) |
| Residential address | SAR (%) | Household | 19.3 (15.5, 23.9) | 23.4 (18.4, 29.3) | 16.4 (13.1, 20.3) | 17.8 (13.8, 22.6) |
|  |  | Non-household | 5.3 (3.6, 7.8) | 6.6 (4.4, 9.8) | 4.7 (3.2, 6.8) | 5.1 (3.4, 7.6) |
|  | Local R | With quarantine | 0.43 (0.35, 0.53) | 0.44 (0.35, 0.55) | 0.43 (0.35, 0.53) | 0.43 (0.34, 0.55) |
|  |  | No quarantine | 0.54 (0.44, 0.67) | 0.66 (0.52, 0.84) | 0.46 (0.37, 0.57) | 0.51 (0.40, 0.64) |

**Table 3.** Model-based estimates (and 95% confidence intervals) of daily transmission probabilities for household contacts 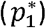 and non-household contacts 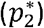 during the incubation and illness periods. Estimates of daily probability of infection from an external source (*b*) and the odds ratios for the relative infectivity during the illness versus incubation period are also provided. Estimates are reported using two different definitions of household contact (close relatives, or only individuals sharing the same residential address) and for selected settings of the natural history of disease. This model is not adjusted for age group.

| Definition of household contact | Parameter | Segment of Infectious Period | Mean incubation period = 4 days |  | Mean incubation period = 7 days |  |
| --- | --- | --- | --- | --- | --- | --- |
|  |  |  | Max infectious period = 13 days | Max infectious period = 19 days | Max infectious period = 13 days | Max infectious period = 19 days |
| Close relatives | $p_1^* (\times 10^{-2})$ | Incubation | 1.58 (1.10, 2.25) | 1.69 (1.21, 2.35) | 2.16 (1.68, 2.77) | 2.18 (1.72, 2.77) |
|  |  | Illness | 1.78 (1.19, 2.66) | 1.34 (0.89, 2.03) | 0.56 (0.20, 1.60) | 0.44 (0.17, 1.12) |
| | $p_2^* (\times 10^{-2})$ | Incubation | 0.79 (0.49, 1.27) | 0.87 (0.55, 1.36) | 1.11 (0.73, 1.67) | 1.13 (0.76, 1.69) |
|  |  | Illness | 0.89 (0.51, 1.55) | 0.69 (0.39, 1.22) | 0.29 (0.09, 0.89) | 0.22 (0.08, 0.64) |
| | $b (\times 10^{-4})$ | | 2.23 (1.14, 4.34) | 1.95 (0.95, 4.01) | 1.53 (0.61, 3.86) | 1.40 (0.54, 3.60) |
|  | Odds Ratio |  | 1.13 (0.59, 2.18) | 0.79 (0.42, 1.49) | 0.26 (0.08, 0.84) | 0.20 (0.07, 0.57) |
| Residential address | $p_1^* (\times 10^{-2})$ | Incubation | 2.40 (1.70, 3.39) | 2.56 (1.85, 3.53) | 3.13 (2.45, 4.01) | 3.16 (2.49, 4.01) |
|  |  | Illness | 2.49 (1.63, 3.79) | 1.89 (1.23, 2.90) | 0.77 (0.26, 2.22) | 0.60 (0.23, 1.56) |
| | $p_2^* (\times 10^{-2})$ | Incubation | 0.62 (0.39, 0.96) | 0.66 (0.43, 1.01) | 0.85 (0.58, 1.23) | 0.86 (0.59, 1.24) |
|  |  | Illness | 0.64 (0.37, 1.11) | 0.49 (0.28, 0.85) | 0.20 (0.07, 0.64) | 0.16 (0.06, 0.45) |
| | $b (\times 10^{-4})$ | | 2.24 (1.14, 4.37) | 1.96 (0.96, 4.03) | 1.52 (0.59, 3.88) | 1.39 (0.54, 3.63) |
|  | Odds Ratio |  | 1.04 (0.54, 2.00) | 0.73 (0.39, 1.39) | 0.24 (0.07, 0.79) | 0.18 (0.06, 0.54) |

Individuals ≥60 years old were the most susceptible group to COVID-19 (Table 4). In comparison to the elderly group (≥60 years old), the odds of infection is 0.27 (0.13-0.55) in the youngest group (<20 years old) and 0.8 (95% CI: 0.54-1.20) in the younger adult group (20-59 years old). These age effects on susceptibility are insensitive to the assumptions about the natural history of disease. Cases <60 years old were slightly less infectious than those ≥60 years old, with ORs varying in the range of 0.73-0.79 but not significantly different from 1 (Table 4; Appendix Table S6). The ORs for the relative infectivity of the incubation vs. illness periods are similar to the model not adjusted for age group. The estimated daily transmission probabilities in the age-adjusted model are for the elderly and therefore higher than the estimates unadjusted for age (Appendix Table S4).

**Table 4.** Model-based odds ratios (and 95% confidence intervals) for age effects on susceptibility and infectivity and relative infectivity during the illness period in comparison to the incubation period. Estimates are reported using two different definitions of household contact (close relatives, or only individuals sharing the same residential address) and for selected settings of the natural history of disease.

| Definition of household contact | Parameter | Odd ratio | Mean incubation period = 4 days |  | Mean incubation period = 7 days |  |
| --- | --- | --- | --- | --- | --- | --- |
|  |  |  | Max infectious period = 13 days | Max infectious period = 19 days | Max infectious period = 13 days | Max infectious period = 19 days |
| Close relatives | Susceptibility | Age group <20 vs. ≥60 | 0.27 (0.13, 0.55) | 0.27 (0.13, 0.54) | 0.26 (0.13, 0.54) | 0.26 (0.13, 0.54) |
|  |  | Age group 20-59 vs. ≥60 | 0.80 (0.53, 1.19) | 0.79 (0.53, 1.18) | 0.81 (0.54, 1.21) | 0.81 (0.54, 1.21) |
|  | Infectivity | Age group <60 vs. vs. ≥60 | 0.79 (0.51, 1.23) | 0.79 (0.52, 1.22) | 0.73 (0.48, 1.12) | 0.73 (0.48, 1.11) |
|  |  | Illness vs. Incubation | 1.14 (0.59, 2.21) | 0.79 (0.42, 1.49) | 0.27 (0.085, 0.82) | 0.19 (0.069, 0.55) |
| Residential address | Susceptibility | Age group <20 vs. ≥60 | 0.23 (0.11, 0.47) | 0.23 (0.11, 0.47) | 0.22 (0.11, 0.46) | 0.22 (0.11, 0.46) |
|  |  | Age group 20-59 vs. ≥60 | 0.74 (0.49, 1.10) | 0.74 (0.49, 1.10) | 0.75 (0.50, 1.12) | 0.75 (0.50, 1.12) |
|  | Infectivity | Age group <60 vs. vs. ≥60 | 0.97 (0.61, 1.54) | 0.97 (0.62, 1.52) | 0.87 (0.56, 1.35) | 0.87 (0.56, 1.34) |
|  |  | Illness vs. Incubation | 1.03 (0.53, 1.98) | 0.72 (0.38, 1.36) | 0.25 (0.10, 0.64) | 0.18 (0.063, 0.52) |

Restricting household contacts to those who were living at the same address as the primary case led to higher SAR estimates among household contacts, 16.4-23.4%, but lower SAR estimates among the non-household contacts under the various settings of the natural history of disease (Table 2; Appendix Table S3). Under the setting of the shortest incubation and infectious periods, the SAR was estimated as 19.3% (95% CI: 15.51-23.86%). The age effects and the relative infectivity of the illness period vs. the incubation period remained qualitatively similar (Table 4; Appendix Table S6).

## Discussion

We characterized the spatial and temporal epidemiology of the COVID-19 epidemic in Guangzhou, the most populated city in southern China. We assessed several aspects of the transmissibility of the disease, such as the time-varying effective reproductive number at the population level, household and non-household SARs, and local reproductive number among close contacts. In addition, we assessed the relative infectivity of the incubation period in comparison to the illness period of COVID-19 cases, and explored the role of age in susceptibility and infectivity. We found that COVID-19 patients in their incubation period were at least as infectious as during their illness period, and that that elderly people are the most susceptible to infection by the SARS-CoV-2 virus, not just severe disease.

When household contact is defined by living at the same address, our model-based household SAR estimate for the Guangzhou contact-tracing data is 19.3%, which is higher than the non-model-based estimates 14.9% for the Shenzhen and 10.2% for Guangzhou.^13,14^ These non-model-based estimates are non-primary attack rates rather than the actual household SARs. In general, a non-primary attack rate is higher than an SAR as the former does not exclude tertiary transmission and untraced exposure. However, these non-primary attack rate estimates reflect transmissibility under control measures such as case isolation, whereas our SAR estimate assumes exposure of a susceptible to the full infectious period of an infector, which is more epidemiologically relevant and generalizable.

Model-based household SAR estimates for the severe acute respiratory syndrome-related coronavirus (SARS-CoV) or the Middle East respiratory syndrome-related coronavirus (MERS-CoV) are not available, but a few studies have reported non-primary attack rates in the household or comparable settings. For SARS-CoV, the non-primary attack rate was estimated to be 4.6% (95%CI: 2.3-8.9%) in Beijing, 8% (95% CI: 6.9-9.1%) in Hong Kong, 6.2% (95% CI: 3.9-8.6%) in Singapore, and 10.2% (95%CI: 6.7-23.5%) in Toronto.^21-23^ Information on household transmissibility of MERS-CoV is less clear. A multi-city household study in Saudi Arabia found the non-primary attack rate to be 4% (95% CI:2-7%).^24^ In an outbreak among female workers who lived in expatriate dormitory consisting of 24 villas in Riyadh, Saudi Arabia, 19 out of 828 were infected. If each villa (24-50 inhabitants) is considered a household, we estimated the non-primary attack rate to be 5.1%.^25^ We conclude that SARS-CoV-2 is more transmissible than both SARS-CoV and MERS-CoV in households.

We are the first to quantify the infectivity of COVID-19 patients during their incubation period using contact-tracing data, but qualitative evidence has been noted before. Transmission to secondary cases during the incubation period of the primary case has been noted in both Germany and China.^6, 26^ In our unpublished analysis of a separate dataset of case clusters in China, the mean serial interval (time between symptom onsets of a primary and a secondary case) was shorter than the mean incubation period, suggesting that infectivity during the incubation period is not trivial.^27^ In contrast, shedding of SARS-CoV peaks 6-11 days after illness onset, and asymptomatic and mild MERS-CoV cases were thought to transmit inefficiently, indicating the key role of symptoms in the transmission of these two coronaviruses.^28-30^ This finding has a profound implication on prevention and control strategies, e.g., the necessity of testing asymptomatic close contacts of COVID-19 cases, and the importance of wearing facial mask regardless of symptoms in order to curb the pandemic.

We estimated the local reproductive number to be relatively low, around 0.48 (Table 3), which is consistent with the average level of *R*_*t*_ (Figure 2). We projected that, without isolation of cases and their contacts, the local reproductive number would have been about 30% higher to reach 0.62 in our primary analysis setting. This moderate effect of isolation is partly due to high infectivity during the incubation period, that is, many secondary cases had been infected before or shortly after the symptom onset of the primary cases before the primary cases were identified and isolated. This observation also implies that the low reproductive number in Guangzhou possibly resulted more from policies restricting human movement, limiting the number of contacts of COVID-19 cases during their entire infectious period including the incubation period, than from the detection and isolation of cases and close contacts.

Our analysis has several limitations. When the mean incubation period is assumed relatively long, the model suggests that the incubation period could be much more infectious than the illness period, which differs from our experience with most respiratory pathogens. This effect could be due to the fact that exposure of close contacts to cases was mostly truncated soon after symptom onset of cases, a consequence of efficient case-finding and isolation of both cases and close contacts all over China after Wuhan’s lockdown. When few transmissions occurred during the illness period, a large bias could happen by chance, as a long incubation period in the model would attribute more transmissions to the incubation period. In addition, we were not able to reliably quantify the infectivity of asymptomatic infections given the limited number of asymptomatic infections, 0.9% and 2.9% among primary and non-primary cases, and our assumption that asymptomatic infections share the same infectivity as the symptomatic cases during their incubation period may not be realistic.

The infectiousness of COVID-19 patients during their incubation period is alarming. This potential of “silent” transmission substantially increased the difficulty in curbing the ongoing pandemic. More and more countries now have realized that active case finding, testing and isolation alone may be inadequate and should be used in conjunction with general human movement restrictions such as stay-at-home policies. On the other hand, the efficient household transmissibility of SARS-CoV-2 should be given full consideration when stay-at-home is ordered. It is crucial to advocate self-monitoring of symptoms and timely self-isolation from other household members when either symptoms emerge or potential exposure occurs. Providing comfortable facilities for exposed contacts to quarantine or for mild cases to isolate away from their families could be a valuable strategy for limiting onward transmission within households.

## Data Availability

Data sharing requests should be sent to Guangzhou Municipal CDC.

## Acknowledgement

This work was supported by grants from the U.S. National Institute of Health (R01 AI139761 and R01 AI116770), the Science and Technology Plan Project of Guangzhou (201804010121), the Project for Key Medicine Discipline Construction of Guangzhou Municipality (2017-2019-04), and the Key research and development program of China (2019YFC1200604). We thank Dr. M. Elizabeth Halloran at Fred Hutchinson Cancer Research Center for helpful discussions. We also thank the staff members of all district-level CDC and community health service centers in Guangzhou for their assistance in field investigation and data collection.

## References

1. Dong E, Du H. and Grdner L. An interactive web-based dashboard to track COVID-19 in real time. Lancet Infectious Diseases. 2020. doi.org/10.1016/S1473-3099(20)30120-1

2. van Doremalen N, Bushmaker T, Morris DH, et al. Aerosol and Surface Stability of SARS-CoV-2 as Compared with SARS-CoV-1. N Engl J Med. 2020. doi.org/10.1056/NEJMc2004973.

3. Xu, Y., Li, X., Zhu, B. et al.. Characteristics of pediatric SARS-CoV-2 infection and potential evidence for persistent fecal viral shedding. Nat Med. 2020. doi.org/10.1038/s41591-020-0817-4.

4. Wu Z and McGoogan, JM. Characteristics of and important lessons from the coronavirus disease 2019 (COVID-19) outbreak in China. JAMA. 2020; : Summary of a Report of 721’314 Cases From the Chinese Center for Disease Control and Prevention. JAMA. 2020. doi:10.1001/jama.2020.2648.

5. Bai Y, Yao L, Wei T, et al. Presumed Asymptomatic Carrier Transmission of COVID-19. JAMA. 2020. doi:10.1001/jama.2020.2565.

6. Rothe C, Schunk M, Sothmann P, et al. Transmission of 2019-nCoV Infection from an Asymptomatic Contact in Germany. N Engl J Med. 2020;382:970-971. doi/full/10.1056/NEJMc2001468.

7. Liu J, Liao X, Qian S, et al. Community transmission of severe acute respiratory syndrome coronavirus 2, Shenzhen, China, 2020. Emerg Infect Dis. 2020. doi.org/10.3201/eid2606.200239.

8. She J, Jiang J, Ye L. et al.. 2019 novel coronavirus of pneumonia in Wuhan, China: emerging attack and management strategies. Clin Trans Med. 2020;9:19. doi.org/10.1186/s40169-020-00271-z.

9. Wu JT, Leung K and Leung GM. Nowcasting and forecasting the potential domestic and international spread of the 2019-nCoV outbreak originating in Wuhan, China: a modelling study. Lancet. 2020. doi.org/10.1016/S0140-6736(20)30260-9.

10. Chinazzi M, Davis JT, Ajelli M, et al. The effect of travel restrictions on the spread of the 2019 novel coronavirus (COVID-19) outbreak. Science. 2020. doi.org/10.1126/science.aba9757.

11. Tian H, Liu Y, Li Y, et al. An investigation of transmission control measures during the first 50 days of the COVID-19 epidemic in China. Science. 2020. doi.org/10.1126/science.abb6105.

12. Liu Y, Eggo RM and Kucharski AJ. Secondary attack rate and superspreading events for SARS-CoV-2. Lancet. 2020. doi.org/10.1016/S0140-6736(20)30462-1.

13. Bi Q, Wu Y, Mei S, et al. Epidemiology and Transmission of COVID-19 in Shenzhen China: Analysis of 391 cases and 1,286 of their close contacts. medRxiv. doi.org/10.1101/2020.03.03.20028423.

14. Luo L, Liu D, Liao X-L, et al. Modes of contact and risk of transmission in COVID-19 among close contacts. medRxiv. doi.org/10.1101/2020.03.24.20042606.

15. Burke RM, Midgley CM, Dratch A, et al. Active Monitoring of Persons Exposed to Patients with Confirmed COVID-19 — United States, January–February 2020. Morbidity and Mortality Weekly Report. 2020; 69:245–246.

16. R Core Team (2017). R: A language and environment for statistical computing. R Foundation for Statistical Computing. Vienna, Austria.

17. Yang Y, Longini IM, Halloran ME and Obenchain V. A hybrid EM and Monte Carlo EM Algorithm and Its Application to Analysis of Transmission of Infectious Diseases. Biometrics. 2012; 68: 1238–1249.

18. Li Q, Guan X, Wu P, et al. Early Transmission Dynamics in Wuhan, China, of Novel Coronavirus-Infected Pneumonia. N Engl J Med. 2020;382:1199–1207.

19. Lauer SA, Grantz KH, Bi Q, et al. The Incubation Period of Coronavirus Disease 2019 (COVID-19) From Publicly Reported Confirmed Cases: Estimation and Application. Ann Intern Med. 2020; doi: https://doi.org/10.7326/M20-0504.

20. Wölfel R, Corman VM, Guggemos W, et al. Virological assessment of hospitalized patients with COVID-2019. Nature (2020). https://doi.org/10.1038/s41586-020-2196-x.

21. Goh DL, Lee BW, Chia KS, et al. Secondary Household Transmission of SARS, Singapore. Emerg Infect Dis. 2004;10: 232–234.

22. Lau JTF, Lau M, Kim JH, et al. Probable Secondary Infections in Households of SARS Patients in Hong Kong. Emerg Infect Dis. 2004;10: 235–243.

23. Wilson-Clark SD, Deeks SL, Gournis E, et al. Household transmission of SARS, 2003. Can Med Assoc J. 2006; 175:1219–1223.

24. Drosten C, Meyer B, Müller MA, et al. Transmission of MERS-Coronavirus in Household Contacts. N Engl J Med 2014; 371:828–835.

25. Van Kerkhove MD, Aswad S, Assiri A, et al. Transmissibility of MERS-CoV infection in closed setting, Riyadh, Saudi Arabia, 2015. Emerg Infect Dis. 2019;25: 1802–1809.

26. Cai J, Sun W, Huang J, Gamber M, Wu J, He G. Indirect virus transmission in cluster of COVID-19 cases, Wenzhou, China, 2020. Emerg Infect Dis. 2020. doi.org/10.3201/eid2606.200412.

27. Lu Q-B, Zhang Y, Liu M-J, et al. Natural history of disease of the novel coronavirus and its implication for infectivity among patients in China. 2020; unpublished.

28. Cheng PK, Wong DA, Tong LK, et al. Viral shedding patterns of coronavirus in patients with probable severe acute respiratory syndrome. Lancet. 2004 May 22;363(9422):1699–700.

29. Memish ZA, Assiri AM and Al-Tawfiq JA. Middle East respiratory syndrome coronavirus (MERS-CoV) viral shedding in the respiratory tract: an observational analysis with infection control implications. Int J Infect Dis. 2014;29:307–8. doi: 10.1016/j.ijid.2014.10.002.

30. Hosani FIA, Kim L, Khudhair A, et al. Serologic Follow-up of Middle East Respiratory Syndrome Coronavirus Cases and Contacts—Abu Dhabi, United Arab Emirates. Clinical Infectious Diseases. 2019;68(3):409–418.

